# Disclosing the true impact of screening endoscopy on colorectal cancer incidence

**DOI:** 10.1101/2022.11.22.22282622

**Authors:** Thomas Heisser, Carlo Senore, Michael Hoffmeister, Lina Jansen, Hermann Brenner

## Abstract

**Objectives:** Randomized trials have demonstrated reduction of colorectal cancer (CRC) incidence by screening endoscopy. However, measured reduction underestimates true reduction due to inclusion of preclinical cases already present at recruitment. We aimed to quantify the true impact of screening endoscopy on reducing the CRC incidence.

**Design:** Simulation study replicating reported CRC incidence by SCORE, a large, randomized screening sigmoidoscopy trial, and deriving expected incidence after excluding cases that manifested during follow-up but were already prevalent at baseline.

**Setting:** Offer of a single flexible sigmoidoscopy in an organised, population-based screening setting.

**Participants:** Simulated, sex- and age-matched SCORE trial population (intervention group, N=17,136, control group, N=17,136, 50% women, ages 55-64 at baseline).

**Interventions:** Screening flexible sigmoidoscopy versus no screening.

**Main outcome measure:** ‘True’ (i.e., unbiased, excluding prevalent cancers at baseline) and ‘apparent’ (i.e., as reported) incidence rate ratios (IRR) for screening versus no screening.

**Results:** In the initial years after randomization, apparent cumulative incidence in the screening group was higher than in the control group due to inclusion of a large proportion of prevalent cancers. In the longer run, apparent cumulative incidence was lower in the screening group, but this incidence reduction was still much lower than true incidence reduction due to inclusion of prevalent cases in calculation of apparent cumulative incidence. In intention-to-screen analysis, apparent/true risk reductions after 8, 11 and 15 years of follow-up were 16%/31%, 20%/28%, and 21/25%, respectively. In per-protocol analyses, respective apparent/true risk reductions were 28%/54%, 34%/49%, and 35%/44%. Estimated underestimation of true incidence was similar among men and women and among age groups 55-59 and 60-64.

**Conclusions:** The preventive effect of screening endoscopy is likely much stronger than reflected in the reported apparent IRRs. Published findings of randomized screening trials underestimate the true preventive effective of screening endoscopy even after 15 year or longer follow-up.

**Summary Box:** *What is already known on this topic:*

- Several large-scale randomized trials have demonstrated substantial reduction of colorectal cancer incidence by endoscopic screening.
- In these trials, the preventive effect of screening endoscopy only transpires after 4-6 years, as screen-detected, prevalent cancers (which can no longer be prevented) dominate the measured incidence in the first years of follow-up.
- The true impact of screening endoscopy on CRC incidence is therefore essentially unknown.

*What this study adds:*

- This modelling study derives estimates of the apparent and true impact of screening sigmoidoscopy on reducing the CRC incidence by accounting for prevalent preclinical cancers at baseline.
- After careful calibration, the model closely predicts observed effects on CRC incidence in the SCORE trial, a randomized trial of flexible sigmoidoscopy conducted in Italy, and demonstrates that the endoscopy screening effect on incidence might be substantially larger when accounting for prevalent preclinical cancers at baseline.

## Introduction

Randomized trials, cohort and modelling studies have consistently demonstrated a major impact of screening endoscopies (flexible sigmoidoscopy or colonoscopy) on reducing colorectal cancer (CRC) incidence and mortality [1,2]. CRC mortality starts to be lower in the screening group compared to the control group from the beginning as a result of earlier detection of prevalent, preclinical (asymptomatic) cases and lowering incidence through removal of precancerous lesions. By contrast, measured incidence shows an initial apparent increase in the screening group due to detection of prevalent, preclinical cancer. Only in the long run, typically after around 4 to 6 years of follow-up, measured cumulative incidence also starts to be lower in the screening group due to later manifestation of initially preclinical cases in the control group and removal of precancerous lesions in the screening group [3–8].

However, the measured incidence rates do not reflect true incidence rates, neither in the screening group nor in the control group, as they in fact are a mix of truly incident cases and cases that were already prevalent in preclinical stage at baseline. The relative share of both case groups in calculation of cumulative incidence strongly depends on the length of follow-up. As screening endoscopies cannot prevent prevalent CRC cases, the commonly measured and reported effects on CRC incidence do not quantify the true endoscopy impact on CRC incidence, i.e., the impact on preventing newly developing CRC cancer.

The aim of this study was to quantify the expected true impact of screening endoscopy on CRC incidence and its expected underestimation by relying on measured incidence rates. We re-calibrated the <u>Co</u>lorectal Cancer Multistate <u>Si</u>mulation <u>Mo</u>del (COSIMO), a previously developed and thoroughly validated modelling approach [9], to reproduce outcomes of the Screening for COlon REctum (SCORE) trial, which provides randomized evidence on the effects of screening by flexible sigmoidoscopy. Then, we derived the expected true incidence in the screening and control group and the difference between the apparent incident rate ratio (IRR_APP_), i.e., the IRR not adjusted for prevalent CRC cases at baseline, and the true incidence rate ratio (IRR_TRUE_), i.e., adjusted for prevalent CRC cases, which is not directly observable in real-life studies.

## Methods

### Multistate Markov Model

We used COSIMO, a previously developed simulation model validated for the German population [9] for our simulations. Briefly, COSIMO simulates the natural history of CRC based on the incidence and progression of precursor lesions developing into preclinical and then clinical cancer, and potential inference by interventions as outlined in detail elsewhere [9] and visually summarized in **Supplementary Figure 1**.

The model’s natural history assumptions were derived step-by-step in several previous analyses using data from the German screening colonoscopy registry, the world’s largest registry of its kind [10–12]. Death rates from CRC were estimated using data from a large population-based case-control study with long-term follow-up of CRC cases and registry data from Germany as previously described [13,14]. General mortality rates and average life expectancy were extracted from German population life tables [15] for the base case model, and from Italian population life tables for simulating the SCORE trial [16]. Sex-specific baseline neoplasm prevalences for each age of screening were extracted from a previous analysis of more than 4.4 million screening colonoscopies in the German-screening eligible population [17].

A comprehensive documentation of the model’s structure and data sources used for its development is given in **Supplementary Appendix 1**. Overviews of key model parameters are provided in **Supplementary Tables 1-3**. The model source code, developed in the statistical software R (version 4.0.2), is available for download from our website [18].

### Simulations

COSIMO was calibrated to simulate the Italian SCORE trial, a large, randomized trial examining the effect of a single flexible sigmoidoscopy in reducing colorectal cancer incidence and mortality (N=34,292). Details on the calibration process are provided in **Supplementary Appendix 2**. In brief, we matched numbers of simulated subjects and allocation per group with reported baseline numbers for SCORE by sex and age [19]. To reflect the not-screen related use of lower gastrointestinal endoscopy outside the study setting (‘contamination’), we assumed an annual colonoscopy use of 0.5-2.0% in the trial population based on data from the European Health Interview Survey (EHIS) for Italy on the years 2013-2016 [20]. Assumptions on referral and surveillance colonoscopies were based on recommendations during the SCORE trial period [19].

### Outcome measures

#### Simulated SCORE: Validation targets

The primary validation objective was to assess the agreement between sex- and age-specific modelled and observed IRR_APPs_ for invited to screening vs control groups (intention-to-screen analysis), as well as for screened vs control groups (per-protocol analysis) over 15 years of follow-up. We therefore calculated model-based IRR_APPs_ by deriving sex- and age-specific numbers of incident cases for intervention and control groups, calculating the incidence rate as number of cases per number of patient-years for each group, and finally calculating the ratio of incidence rates of the intervention (screened) group and the control group. Outcomes were compared to those extracted for the actual SCORE trial [6]. Results were considered consistent if modelled estimates were within the 95%-confidence intervals (CI) of the corresponding outcomes reported for the SCORE trial.

#### Apparent vs true incidence

Subsequently, to determine the impact of prevalent preclinical cancers on reported IRR_APP_, we calculated the IRR_TRUE_ for intervention vs control groups by omitting the sex- and age-specific number of incident cases arising from prevalent preclinical cancers (CRC_preclin_) from the model calculation of IRRs for each year of follow-up. We also calculated the corresponding ‘true’ cumulative incidence for each group over time and compared the respective outcomes. Differences between IRR_APP_ and IRR_TRUE_ were expressed by calculating the absolute and relative underestimation of the estimated incidence reduction by screening.

### Patient and Public Involvement

Patients and the public were neither involved in the design and conduct of this study, nor in writing or editing of this document. Research at the German Cancer Research Center (DKFZ) is generally informed by a Patient Advisory Committee.

## Results

### Validation of reproduced SCORE

**Table 1** shows the number of incident cases, the incidence rate as well as IRRs by sex and age for those invited to screening compared to the control group as reported for the actual and simulated SCORE trial after 15 years of follow-up. Incidence rates predicted by COSIMO were very similar to those observed for the actual SCORE trial. COSIMO predicted 207 and 117 cases per 100,000 men and women in the intervention group, and 260 and 146 cases per 100,000 men and women in the control group, resulting in IRR_APPs_ of 0.79 for men and 0.80 for women, consistent with the SCORE results **(Supplementary Figure 2A)**. Modeled and actual age-specific IRR_APPs_ were likewise similar and close to 0.8 in each case **(Supplementary Figure 2B)**. All primary validation targets were reached since the modelled IRR_APP_ point estimates were within the reported 95% CIs of the actual trial.

**Table 1.**
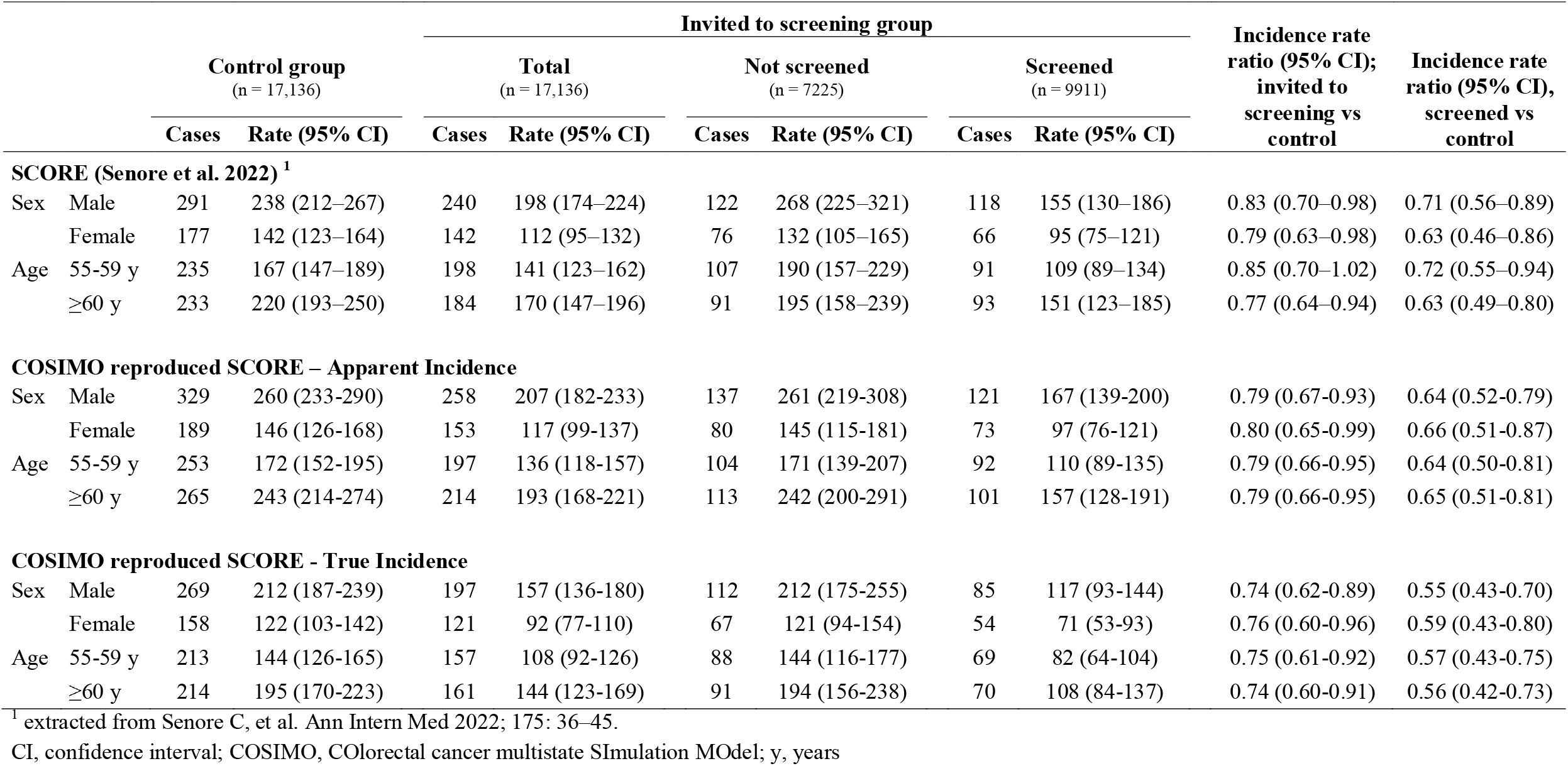
Reported and simulated apparent and true colorectal cancer incidence in the SCORE trial by sex and age groups, randomization and compliance with screening after 15 years of follow-up.

Over time, the (apparent) cumulative incidence in the simulated screening group increased markedly in the year following the screening, followed by modest further increase until the end of follow-up (**Supplementary Figure 3**). In the control group, the (apparent) cumulative incidence followed a steady growth trajectory. Both curves crossed after approximately 5 years, with the gap between curves widening up with longer follow-up duration, a pattern also observed in the actual SCORE trial.

### True Incidence

Excluding prevalent preclinical cancers at baseline markedly changed the number of detected CRC cases as well as incidence rates. The relative share of prevalent, screen-detected cases among all detected cases was higher in the intervention than in the control group but exceeded 50% in the initial years of follow-up in both groups (**Figure 1**). The share of prevalent cases from all cumulatively reported cases diminished with increasing length of follow-up but even after 15 years of follow-up was still as high as 22.5% and 17.5% in the intervention and control group, respectively.

**Figure 1.**
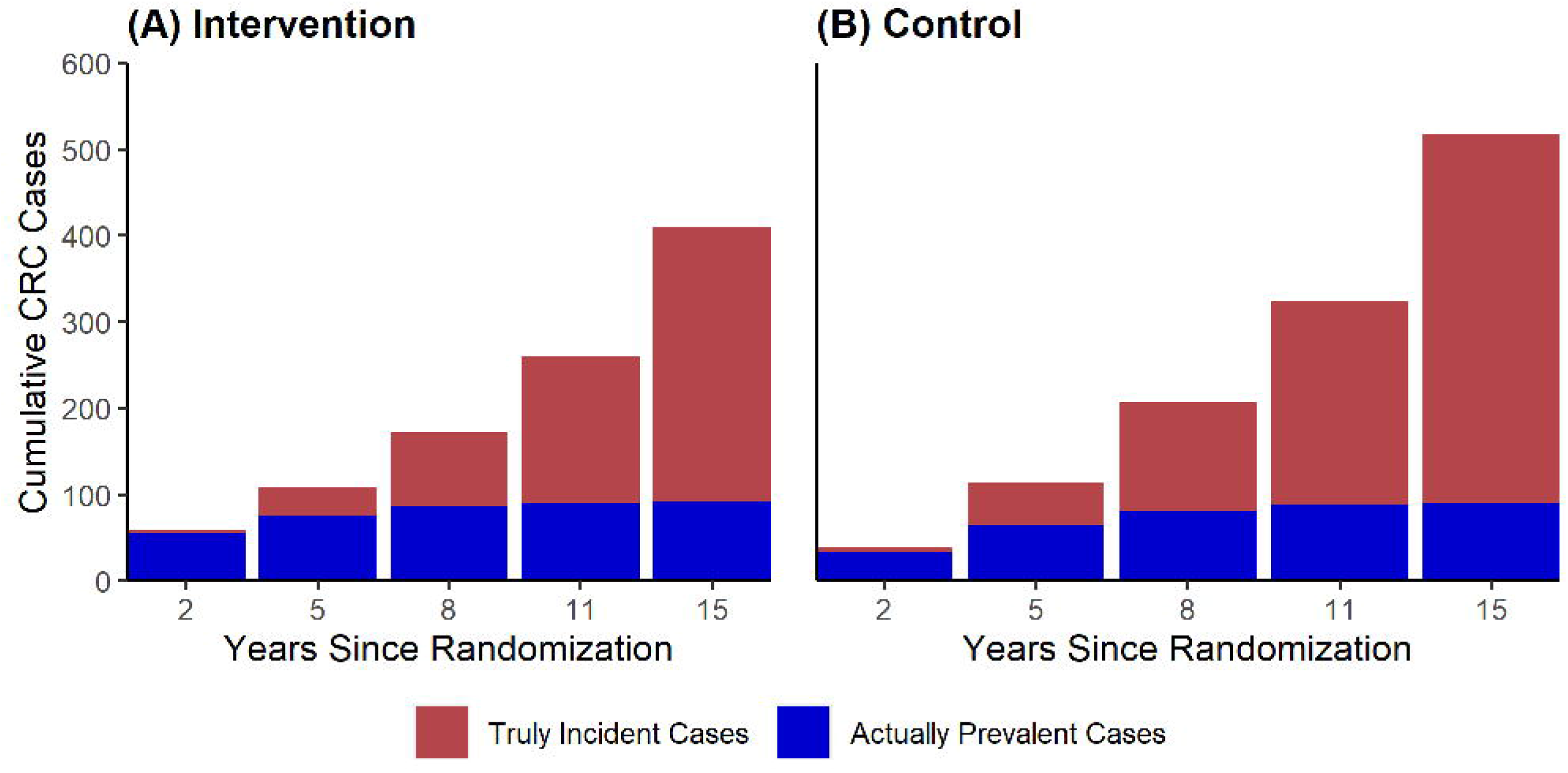
Combined numbers of truly incident as well as prevalent, screen-detected colorectal cancer cases in the simulated SCORE trial over time. CRC, colorectal cancer

**Table 2** provides estimates of the apparent and true IRRs (calculated with and without inclusion of prevalent cases) according to year of follow-up. Even though both IRR_APP_ and IRR_TRUE_ were lower than 1, indicating a protective effect of screening, after five or more years of follow-up, and the difference between IRR_APP_ and IRR_TRUE_ diminished with increasing length of follow-up, true incidence reduction was still underestimated by 16% and 20% in intention-to-screen and per-protocol analyses, respectively, even after 15 years of follow-up (**Figure 2)**. This pattern of notably stronger true than apparent protective effects of screening sigmoidoscopy was consistently found among men and women (**Supplementary Table 5, Supplementary Figures 4 and 5)** and seen for both younger (55-59 years) and older (60-64 years) study participants **(Supplementary Table 6, Supplementary Figures 6 and 7)**.

**Figure 2.**
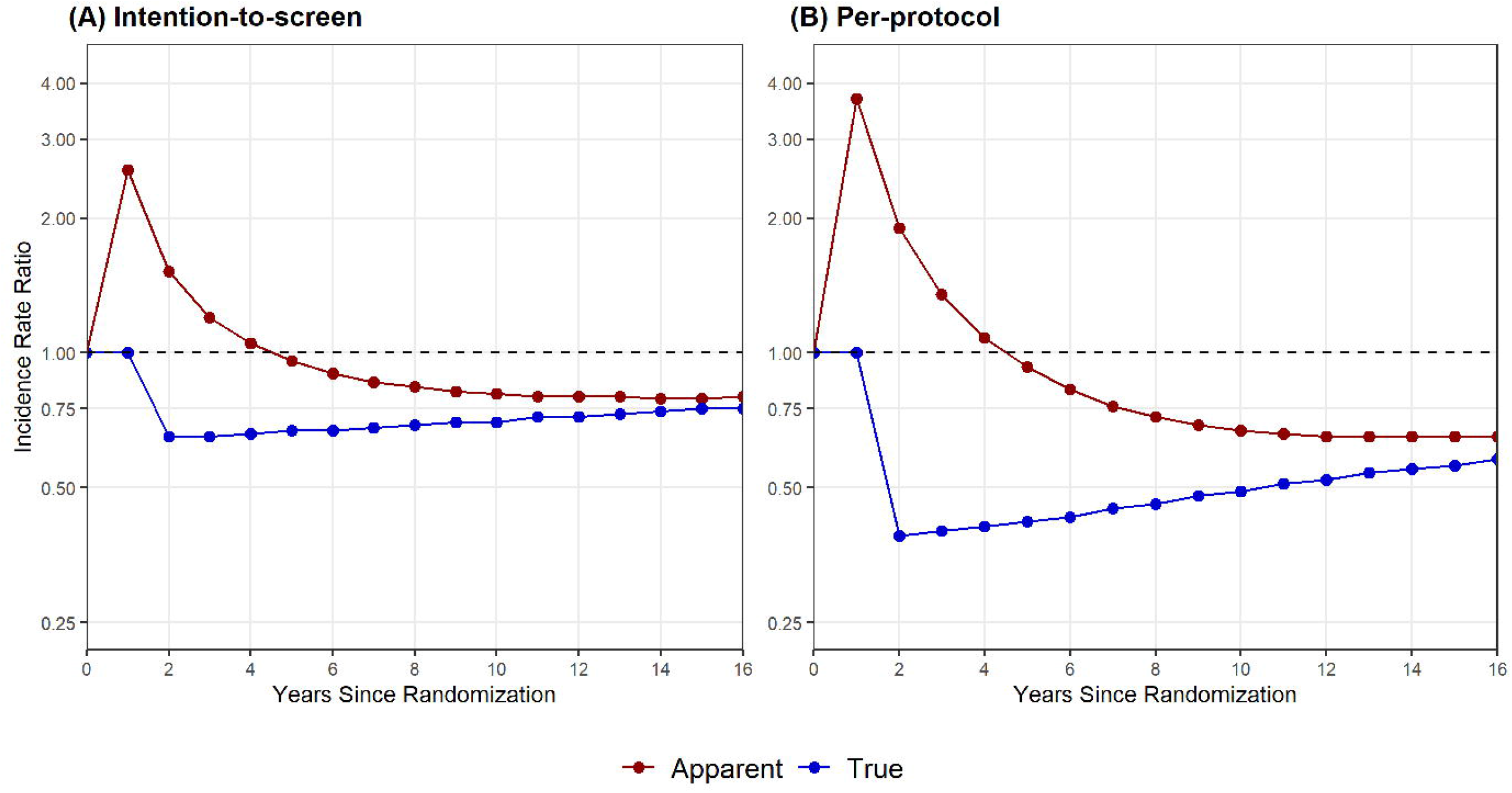
Apparent and true incidence rate ratios in the simulated SCORE trial over time.

**Table 2.** Apparent and true incidence rate ratios, risk reduction and underestimation of risk reduction in the simulated SCORE trial over time.

| Year of follow-up | IRR |  | Risk reduction |  | Underestimation of risk reduction |  |
| --- | --- | --- | --- | --- | --- | --- |
|  | Apparent | True<sup>1</sup> | Apparent | True<sup>1</sup> | Absolute | Relative |
| **Intention-to-screen analysis** |  |  |  |  |  |  |
| 5 | 0.96 | 0.67 | 4% | 33% | 29 % units | 88% |
| 8 | 0.84 | 0.69 | 16% | 31% | 15 % units | 48% |
| 11 | 0.80 | 0.72 | 20% | 28% | 8 % units | 29% |
| 15 | 0.79 | 0.75 | 21% | 25% | 4 % units | 16% |
| **Per-protocol analysis** |  |  |  |  |  |  |
| 5 | 0.93 | 0.42 | 7% | 58% | 51 % units | 88% |
| 8 | 0.72 | 0.46 | 28% | 54% | 26 % units | 48% |
| 11 | 0.66 | 0.51 | 34% | 49% | 15 % units | 31% |
| 15 | 0.65 | 0.56 | 35% | 44% | 9 % units | 20% |

<sup>1</sup> 'true' incidence rate ratio / risk reduction: excluding prevalent cancers at baseline not preventable by screening.
IRR, incidence rate ratio

Finally, in line with these findings, the ‘true’ cumulative incidence was consistently lower than the ‘apparent’ cumulative incidence, without the characteristic overlapping of curves after 5 years in graphical analyses **(Figure 3)**. Instead, the curves started widening up already starting from baseline, illustrating that the intervention (screened) groups were favored throughout the entire observation period.

**Figure 3.**
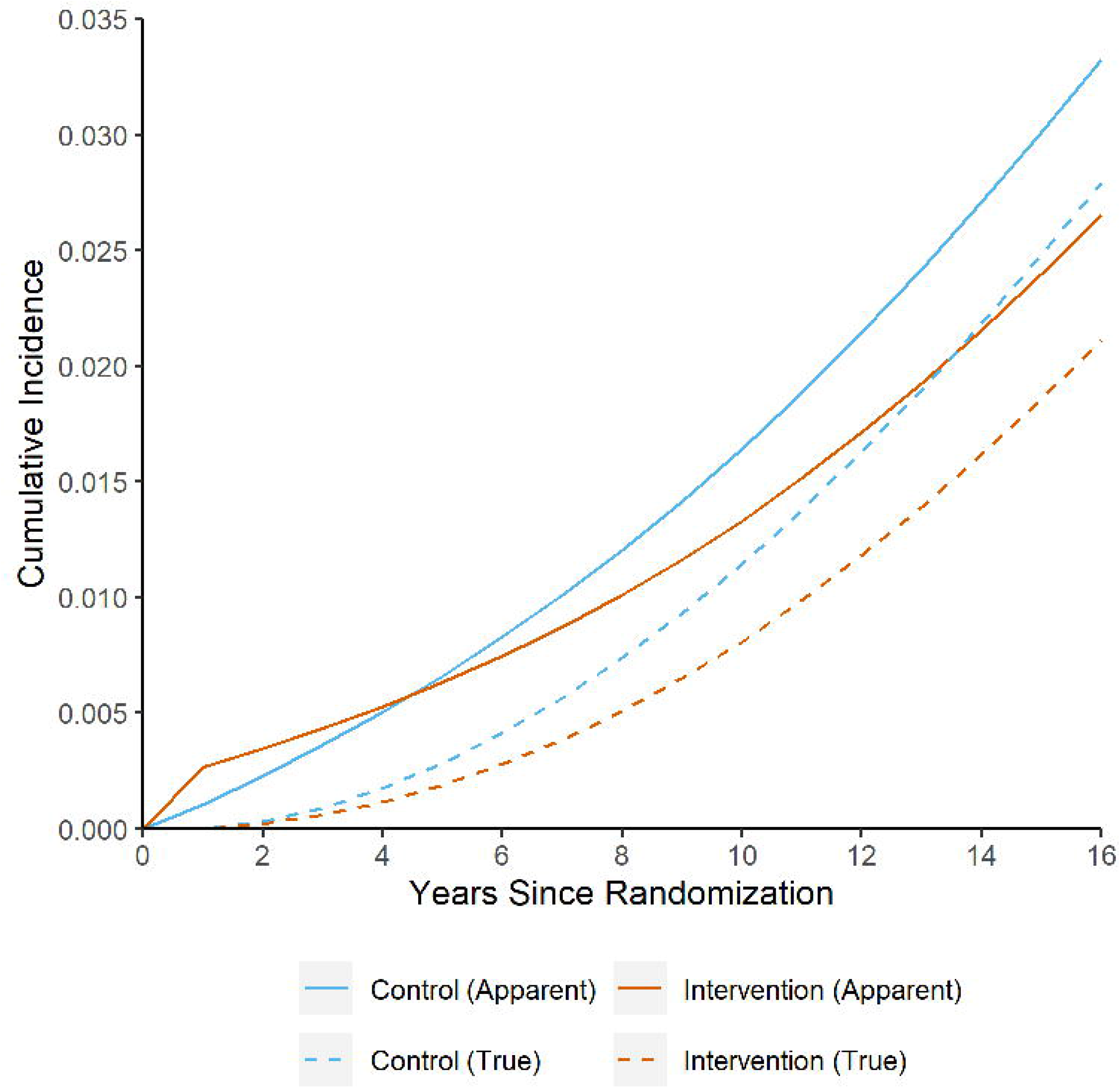
Apparent and true cumulative colorectal cancer incidence (intention-to-screen analysis) in the simulated SCORE trial over time.

## Discussion

Here, we illustrated the underestimation of the true impact of screening sigmoidoscopy on reducing the CRC incidence. We first replicated the reported CRC incidence in SCORE, a large, randomized screening sigmoidoscopy trial, and then derived the expected incidence after excluding cases that manifested during follow-up but were already prevalent at baseline. We found that, in the initial years after randomization, apparent cumulative incidence was higher in the screening versus control group, reflecting the inclusion of a large proportion of prevalent cancers. Only with increasing length of follow-up, apparent cumulative incidence started to be lower in the screening group. However, even after 15 years, apparent incidence reduction was still much lower than the ‘true’ (i.e., unbiased) incidence reduction, as prevalent cases were still included in the calculation of apparent cumulative incidence. Taken together, these results suggest a much stronger and much earlier preventive effect of screening sigmoidoscopy than reflected in the reported apparent incidence rate ratios.

### Findings in Context

The effectiveness of screening sigmoidoscopy to reduce CRC risks has been studied across in total four RCTs, with reported cumulative incidence reductions after median 14-17 years of follow-up ranging from 18-26% in intention-to-screen and 33-35% in per-protocol analysis [3–7]). All RCTs have in common that the preventive effect of screening sigmoidoscopy only transpired after 4-5 years. Though the reason for this behavior (i.e., the dominance of screen-detected prevalent cancers, which could not any longer be prevented, in the first years of follow-up) has been previously noted [21,22], to our knowledge, no previous attempt has been made to quantify the impact of these prevalent cancers, and their relative contribution to the overall reported incidence reduction remained unclear.

By simulating a scenario without prevalent preclinical cancers at baseline, this modelling study adds such quantification of the true impact of endoscopy on CRC incidence reduction to the literature. The findings indicate that the preventive potential of screening endoscopy is likely much larger than previously reported, most notably within the first 10 years after the intervention and still not fully discernable even after 15 years of follow-up. Excluding prevalent cancers had several implications: first, there was no crossing of incidence curves after 4-5 years (characteristic for the sigmoidoscopy RCTs [3–6]), and the intervention group was strongly favored from the beginning. Second, while the IRR_APPs_ tended to improve in favor of the intervention with increasing duration of follow-up, IRR_TRUEs_ suggested the strongest difference between intervention vs control early after screening, and more muted (but still strong) differences with increasing follow-up duration. Third, while IRR_TRUEs_ were consistently more favorable towards the intervention than IRR_APPs_, the magnitude of the underestimation strongly depended on the time of follow-up (e.g., in intention-to-screen analyses, the ‘true’ incidence reduction by screening was 31 per cent points higher after 5 years, but only 4 per cent points higher after 15 years), illustrating the diminishing impact of prevalent preclinical cases with increasing duration of follow-up. Fourth, differences between IRR_TRUEs_ and IRR_APPs_ were consistent across sexes as well as age groups.

Regarding the latter, it may be noted that the magnitude of the preventive potential of sigmoidoscopy in women is not undisputed. The evidence from sigmoidoscopy RCTs is ambiguous, indicating a similar or possibly greater (SCORE), smaller (PLCO, UKFSS) or even no (NORCCAP) effect of screening in women. A pooled analysis suggested stronger incidence reduction in younger women versus men but no screening effect in older women [23]. However, the CIs of these sex-specific estimates overlap, and chance findings due to underpowered studies by limited sample size for sex-stratified analyses cannot be ruled out. Considering our findings, a beneficial effect of screening sigmoidoscopy could be hypothesized even for the corresponding female NORCCAP study population, as excluding prevalent preclinical cancers would likely tilt the outcome estimates more in favor of the intervention group. Such analyses should be addressed in future research.

Long-term outcomes from several RCTs on the effects of screening colonoscopy are still pending [24–27]. Initial results from the north-European NordICC trial were recently published [8]. However, given that follow-up so far was limited to 10 years, results are to be considered preliminary [28]. In particular, our results suggest that underestimation of true incidence reduction by inclusion of a high proportion of prevalent cases may still be substantial. The combined body of evidence from case-control, cohort and simulation studies suggests that the preventive potential may even be larger than for sigmoidoscopy [29]. Colonoscopy reaches the entire colon and may thus also detect precursor lesions in the proximal colon with high sensitivity, which may imply higher potential to reduce CRC incidence over time.

The flexible sigmoidoscopy RCTs also showed a significant reduction of CRC mortality. However, while excluding prevalent, preclinical cancers at baseline would most likely lead to lower CRC mortality rates in both intervention and control groups, no substantial impact on corresponding estimates of the relative difference between both arms will be expected. In contrast to CRC incidence (which cannot any longer be lowered by early detection of prevalent cases), screening can also contribute to lowering CRC mortality through early detection of prevalent cancers, as most early-stage cancers can be treated with curative intent, e.g., by surgical removal. The inclusion of prevalent, preclinical cancers at baseline is therefore not to the same extend prone to bias estimates of effects on CRC mortality.

### Limitations

A key limitation of this study is that calibrating COSIMO to replicate SCORE relied on several assumptions for model input parameters. For instance, as the number of patients by age was only published on group-level [6], we assumed a uniform distribution of patients across individual ages in the model. Although this will have introduced an additional source of variability, it will likely not have impacted the outcomes to any meaningful extent, as prevalences and transition rates within these age groups are overall very similar. Other sources of uncertainty include the proportion of screening endoscopy users outside of the trial (‘contamination’), the prevalences of CRC precursor lesions at baseline, and the adenoma miss rate of sigmoidoscopy within SCORE. However, as we used the best available evidence from the literature or performed additional calculations to derive input parameters as approximation, we believe these assumptions to be robust, which is also evidenced by meeting of all validation targets.

A further important limitation of COSIMO is that no distinction according to cancer subsite can be made by the model. This is as the main structural parameters of the model, the transition rates between states, were originally derived from the German national screening colonoscopy registry, the world’s largest of its kind [10–12]. Though the registry represents a particularly well-suited data source to derive such transition rates, it did unfortunately not include sufficiently detailed data to calculate specific rates for proximal and distal neoplasms. We could therefore not provide estimates on the true CRC incidence separately for distal vs proximal colon, which would have been desirable given the preventive potential of screening sigmoidoscopy primarily extends to the distal part of the colon.

Due to limited scope, even though several flexible sigmoidoscopy RCTs have reported results on CRC incidence reduction which likely are likewise strongly underestimated, COSIMO was only calibrated to replicate the Italian SCORE trial. On similar grounds, we abstained from health-economic considerations, which might also be implicated by our findings. However, we would expect similar patterns of underestimation of true incidence reductions for the other flexible sigmoidoscopy RCTs, and perhaps even stronger underestimation for the NORDICC colonoscopy trial due to the even larger proportion of prevalent cancers detected by screening colonoscopy which should be specifically addressed in future studies.

## Conclusions

In randomized trials, the true impact of screening endoscopy on reducing colorectal cancer incidence is partly masked by the inclusion of prevalent, preclinical cancers at baseline which cannot any longer be prevented. The relative share of such prevalent screen-detected cases from all detected cases strongly depends on the length of follow-up and diminishes over time. Excluding prevalent preclinical cancers at baseline in a replicated, simulated version of the randomized SCORE trial suggests that the true incidence reduction by screening sigmoidoscopy is strongly underestimated in the first 10-years of follow-up and still underestimated by approximately 16-20% even after 15 years as compared to the corresponding published estimates. Thus, the preventive effect of screening endoscopy is likely much stronger, and manifests much earlier than previously reported. Published findings of randomized screening trials significantly underestimate the true preventive effective of screening endoscopy.

## Data Availability

All analyses relevant to the study are included in the article or uploaded as supplementary information. The model source code is available from https://www.dkfz.de/en/klinepi/download/index.html. Further information is available from the corresponding author upon request

