## Supplementary Material for "Disclosing the true impact of screening endoscopy on colorectal cancer incidence"

True Colorectal Cancer Incidence

Thomas Heisser, Carlo Senore, Michael Hoffmeister, Lina Jansen, Hermann Brenner

### **Supplementary Appendix 1. COSIMO Model Documentation**

#### **Conceptual model structure**

Our Markov-based Colorectal Cancer Multistate Simulation Model (COSIMO) simulates the natural history of CRC based on the process of precursor lesions (non-advanced and advanced adenomas) developing into preclinical (asymptomatic) and then clinical (symptomatic) cancer. In the core model, the simulation is performed on a hypothetical previously unscreened German population, with the number of simulated subjects and their corresponding baseline age (minimum 50 years) being variables to be chosen prior to model start. For this project, COSIMO was updated to simulate the SCORE trial, which was conducted in an Italian population.

At start of the simulation, certain proportions of no neoplasm, non-advanced adenoma, advanced adenoma and preclinical CRC are assigned to the hypothetical population. The simulation runs up to a predefined number of cycles of each one year. Each year, people at each state have a certain probability (transition rate) to progress to the next state. Subjects with CRC may die from the disease, and at each state people may experience non-CRC death, reflecting the general background mortality from other causes.

Screening can alter the progression between states. People with adenoma will be moved backward to the state of no neoplasm, assuming removal of their adenoma at colonoscopy (for screening or diagnostic workup, e.g., after a positive fecal test). Subjects will then continue to have the probabilities to progress to the next states as those without findings at screening. We assume that, although these people are under a higher risk of developing adenomas or cancers than the general population [1], the excess risk will be effectively compensated through the protection provided by surveillance colonoscopies [2,3]. Preclinical CRC detected at screening will be moved forward to the state of diagnosed cancer.

After each cycle where a screening test was applied, the model differentiates the simulated population into a ‘screening negative’ and a ‘screening positive’ group, which allows for modelling different trajectories depending on the screening outcome. This feature enables the implementation of more complex screening and surveillance strategies and scenarios. For instance, it facilitates modelling scenarios where subjects only receive the next screening round if they had a negative test result in the respective previous round (e.g., in stool test screening). Also, subjects with false-positive test results may return to the screening population after a predefined latency period (e.g., 10 years for screening colonoscopy scenarios). Finally, it allows the implementation of surveillance after true-positive test results. In the base case model, subjects with detected non-advanced adenomas are assumed to undergo surveillance colonoscopies at intervals of 10 years up to a predefined end age of 75 years. In case an advanced adenoma was detected, either at the primary screening test or at a surveillance colonoscopy, subjects are assumed to undergo periodic surveillance colonoscopies at three-yearly intervals up a predefined end age of 85.

The model source code, developed in the statistical software R (version 4.0.2), is available for download from our website [4].

#### **Model parameters**

**Starting prevalences and transition rates of the core model**

An overview of key model parameters is given in **Supplementary** **Table 1**.

*Data source*

The data basis of our analyses on model starting prevalences and transition rates was the nationwide screening colonoscopy registry run by the Central Research Institute of Ambulatory Health Care in Germany. The registry, which was built up along with the introduction of the screening colonoscopy offer in the year 2002, is a repository of all screening colonoscopies conducted in Germany. Reporting is virtually complete, as it is a prerequisite for physicians’ reimbursement by the health insurance funds. The registry includes only primary screening examinations (i.e., colonoscopies conducted for surveillance, work-up of symptoms or other screening tests are not included). Items reported include, besides basic sociodemographic variables, findings at colonoscopy, including number, size and histological characteristics of polyps. In case of multiple neoplasms, only the most advanced one (non-advanced adenoma, advanced adenoma, or cancer) is recorded. Advanced adenomas are defined as at least 1 adenoma ≥ 1 cm or at least 1 adenoma with villous components or high-grade dysplasia.

Noteworthy, the reporting for the screening colonoscopy registry does not differentiate by the class of lesion. Thus, the herein used term ‘adenoma’ refers to conventional or serrated adenomas (polyps) alike. While we preferred to refer to our model as being based on the adenoma-carcinoma pathway in previous publications [5–9] for the sake of simplicity and comprehensibility (as the grand majority of CRCs develops through this well-established pathway of cancer development [10,11]), in fact COSIMO’s defining parameters were derived using polyp/adenoma prevalences as detected and reported at screening colonoscopy, regardless of their underlying mechanism or pathway of development. Therefore, it will be more precise to refer to the model as being based on the ‘natural history of CRC’, without restrictions on underlying CRC development pathways.

*Starting prevalence*

The sex- and age-specific proportions of no neoplasm, non-advanced adenoma, advanced adenoma and preclinical CRC at the beginning of simulation were calculated based on the data from 4.4 million participants of the German screening colonoscopy program who had their first screening colonoscopy during 2003–2012 [5]. To take into account that a certain proportion of neoplasms needs to be assumed to have been missed at colonoscopy screening, in particular for serrated or flat polyps [12,13], we re-calculated the previously reported prevalences, assuming representative miss rates of 25% for non-advanced adenomas and 5% for advanced neoplasms (advanced adenomas and preclinical cancers).

The prevalences found in those aged 55 were used as the best estimate for simulations starting with a 50-year-old population, which seems reasonable as selected regional programs which offer screening colonoscopy from age 50 on found similar prevalences of adenomas in age groups 50-54 and 55-59 [14].

*Transition rates*

Transition rates between states were estimated based on data from the nationwide screening colonoscopy registry by several separate birth cohort and mean sojourn time analysis. [15,16] Briefly, sex- and age-specific annual incidence and transition rates were estimated from sex- and age-specific prevalences of adenomas among 3.6 – 4.3 million screening participants from the same birth cohorts in 2003–2011 (2003-2009) and 2004–2012 (2004 – 2010) as reported to and documented in the screening colonoscopy registry (see above for details on the data source). The analysis on mean sojourn time of preclinical cancers additionally incorporated registry-reported colorectal cancer incidence and participation rates in screening colonoscopy from 2003-2006.

Similar as for the starting prevalences, as colonoscopy was shown to be less effective in detecting serrated lesions (and as the true proportions of missed conventional adenomas and serrated lesions in the registry-reported prevalences is unknown), we re-calculated previously reported transition rates [15–17] to adjust for representative colonoscopy miss rates [12,13]. This adjustment resulted in slightly higher overall prevalences of adenomas, and therefore (when compared to previously reported rates) in slightly higher transition rates of incidence adenomas, as well as slightly lower transition rates from non-advanced to advanced adenomas and from adenomas to cancer. Furthermore, to adjust for uncertainties resulting from the cycle length of one year used in COSIMO, we updated the model to allow for small proportion of subjects with very rapidly progressing lesions with limited potential for early detection and associated worse prognosis. Age- and sex-specific annual transition rates between the states were estimated for age groups from 55-79 years in steps of 5 years. Estimates for age 50-54 and ≥ 80 (or ≥ 85) were assumed to be the same as those for age group 55-59 and 75-79 (or 80-84), respectively.

Confidence intervals for both starting prevalences and transition rates were derived by bootstrap analysis with resampling within sex- and age-specific subgroups. Ninety-five percent confidence intervals were determined as the 2.5th and 97.5th percentile of transition rate estimates obtained in 1,000 runs.

**Mortality rates**

Mortality rates for patients whose cancer was detected by screening or by symptoms were estimated in previous analyses [8,9]. We combined data on the proportion of screening-detected cases among all CRC cases in Germany during 2003-2012 in people aged 55-79 years [5,18] with the overall CRC-specific mortality rates by year after diagnosis in Germany in 2011-2012 [18]. We then used hazard ratios for patients detected by screening versus symptoms as obtained from a German population-based case-control study on CRC screening with long-term mortality follow-up of CRC patients [8,19] to estimate CRC-specific mortality rates by mode of detection (**Supplementary** **Table 2**). Sex- and age-specific general mortality rates and average life expectancy of the population were extracted from German and Italian population life tables 2010/2012 (**Supplementary** **Table 3)** [20].

**Model validation**

COSIMO has been validated for the German screening-eligible population. Details on the model validation process can be found in the literature [21]. Briefly, we pursued a three-fold approach using the best available evidence from epidemiological data sources in Germany. We compared model-derived cumulative incidence and prevalences of colorectal neoplasms to (a) results from KolosSal, a study in German screening colonoscopy participants, (b) registry-based estimates of CRC incidence in Germany, and (c) outcome patterns of randomized sigmoidoscopy screening studies. This approach enabled us to scrutinize the model's natural history component (Parts a and b) as well as the modeled effect of screening colonoscopy (Parts b and c) at the same time.

We found that (a) more than 90% of observed prevalences in the KolosSal study were within the 95% confidence intervals of the model-predicted neoplasm prevalences; (b) the 15-year cumulative CRC incidences estimated by simulations for the German population deviated by 0.0% to 0.2% units in men and 0.0% to 0.3% units in women when compared to corresponding registry-derived estimates; and (c) the time course of cumulative CRC incidence and mortality in the modeled intervention group and control group closely resembles the time course reported from sigmoidoscopy screening trials. Overall, COSIMO adequately predicted colorectal neoplasm prevalences and incidences in a German population for up to 25 years, with estimated patterns of the effect of screening colonoscopy resembling those seen in registry data and real-world studies.

### **Supplementary Appendix 2. Details on the calibration process**

COSIMO was calibrated to simulate the SCORE trial, a large, randomized trial examining the effect of a single flexible sigmoidoscopy in reducing colorectal cancer incidence and mortality (N=34,292). Details on SCORE have been reported in the literature [22–24]. In brief, participants were recruited from six centers in northern Italy between June 1995 and April 1999. Men and women aged 55-64 and at average risk for CRC were eligible[22]. Interested respondents were assigned randomly to the control group (no further contact) or the intervention group (invitation to undergo a single screening sigmoidoscopy). Screenees with high-risk polyps were referred for colonoscopy. Recently, the authors reported that the strong protective effect of a single sigmoidoscopy screening for CRC incidence and mortality was maintained after a median of follow-up of 15.4 years and 18.8 years, respectively[24].

To reflect trial population and design of a SCORE trial simulated in COSIMO, we matched numbers of simulated subjects and allocation per group with reported baseline numbers for SCORE by sex and age, which were extracted from the most recent publication[24]. Only those invited to screening and screened were assumed to undergo screening sigmoidoscopy at study baseline, and no intervention was assumed for those reported as invited to screening but not screened and controls. For both screened and unscreened groups, separate models were estimated for each sex- and age-specific cohort for 16 years of follow-up, and subsequently combined. As the number of subjects included in SCORE has only been reported in 5-year age-groups (55-59 and 60-64), a uniform distribution across individual ages was assumed.

In addition, as the COSIMO core model had been estimated for the German population, the proportion of preclinical cancers at baseline as well as the incident rate of non-advanced adenomas was discounted by 6% to reflect the lower incident rate in Italy versus the German population, as derived from reported differences in the literature before and shortly after the uptake of population-wide CRC screening offers as a proxy[25] (**Supplementary Table 4**).

*Diagnostic performance of sigmoidoscopy*

COSIMO was built on data from the German screening colonoscopy registry, which unfortunately did not include sufficiently detailed data to calculate specific transition rates for proximal and distal neoplasms. Therefore, COSIMO does not distinguish according to cancer subsite. As the reach of sigmoidoscopy only extends to the distal part of the colon, we derived the expected sensitivity of sigmoidoscopy relative to the entire colon as a surrogate input parameter for COSIMO.

To this end, we assessed the sex-specific proportion of colonoscopy-detected adenomas in the distal part of the colon (rectum, rectosigmoid, and sigmoid colon) compared to all detected colorectal adenomas in the Begleitende Evaluierung innovativer Testverfahren zur Darmkrebs-Früherkennung (BLITZ) study, an ongoing screening study in Germany **(Supplementary Table 1)**. Participants are recruited in 20 gastroenterology practices since end of the year 2005, and, by the end of March 2019, 11,104 participants were recruited. Further details on BLITZ studies have been reported previously [26]. As lesions were detected by colonoscopy in BLITZ, our approximation approach assumes sigmoidoscopy to have the same sensitivity as colonoscopy (within the reach of the sigmoidoscope), in line with previous studies [27,28]. However, the baseline screening in SCORE was conducted between 1995-1999, i.e., up to two decades earlier than in BLITZ. We therefore discounted the BLITZ-derived sensitivity for non-advanced findings by 25% and the sensitivity for advanced findings by 5% to account for substantial improvements in adenoma detection rates over time [29]. Finally, based on the management of SCORE trial participants [22], we assumed detection of proximal neoplasms (if present) in 70% of individuals with distal findings.

*Colonoscopy use outside of study and for surveillance*

To reflect the not-screen related use of lower gastrointestinal endoscopy outside of the study setting (‘contamination’), we assumed an annual colonoscopy use of 0.5-2.0% in the trial population. For SCORE, no data on the degree of such contamination has been reported. However, a study using data from the European Health Interview Survey (EHIS) on the years 2013-2016 reported approximately 22% colonoscopy use within 10 years in the Italian population aged 50-74 [30]. Given the only partly overlapping observation and reporting periods as well as the broader availability and popularity of fecal testing over time [23,31,32], we considered relatively lower annual contamination levels of 0.5-2.0% as an appropriate base case assumption. Contamination was assumed to be slightly higher in the non-screened group (ages 55-59, 1.0%, ages 60-64, 2.0%) versus the screened group (ages 55-59, 0.5%, ages 60-64, 1.0%), and higher contamination in those aged 60-64 versus those aged 55-59 was assumed in line with reports indicating higher levels of endoscopy use with increasing age [33].

Finally, we assumed use of surveillance colonoscopies as recommended for individuals with positive findings at baseline sigmoidoscopy. Subjects with non-advanced adenomas were assumed to undergo surveillance colonoscopies at ten-yearly intervals up to age 75. In case an advanced adenoma was detected, either at screening sigmoidoscopy or surveillance colonoscopy, subjects were assumed to undergo periodic surveillance colonoscopies at three-yearly intervals, reflecting recommendations during the SCORE trial period [22].

### **Supplementary Appendix 3. Supplementary Tables and Figures**

**Overview**

**Supplementary Table 1.** Overview of model parameters

**Supplementary Table 2.** Annual CRC-specific mortality rates of CRC patients by mode of cancer detection

**Supplementary Table 3.** Sex- and age-specific general mortality rates

**Supplementary Table 4**. Standardized colorectal cancer incidence rate in Germany and Italy, 2000-2004

**Supplementary Figure 1**. Schematic illustration of the Colorectal Cancer Multistate Simulation Model (COSIMO)

**Supplementary Figure 2.** Colorectal cancer incidence rate ratios (‘apparent’) in the actual as well as the simulated SCORE trial by sex and age groups after 15 years of follow-up

**Supplementary Figure 3**. Cumulative incidence of colorectal cancer by time from randomization in the actual and simulated SCORE trial (overall trial population, intention-to-screen analysis)

**Supplementary Figure 4.** Apparent and true incidence rate ratios (intention-to-screen analysis) in the simulated SCORE trial over time, stratified by sex

**Supplementary Figure 5.** Apparent and true incidence rate ratios (per-protocol analysis) in the simulated SCORE trial over time, stratified by sex

**Supplementary Figure 6**. Apparent and true incidence rate ratios (intention-to-screen analysis) in the simulated SCORE trial over time, stratified by age

**Supplementary Figure 7.** Apparent and true incidence rate ratios (per-protocol analysis) in the simulated SCORE trial over time, stratified by age

#### **Supplementary Table 1** Overview of model parameters

| **A1. Core model. Proportions of no neoplasm, non-advanced adenoma, advanced adenoma and preclinical CRC at the beginning of simulation**^1^ | | | | | |
| --- | --- | --- | --- | --- | --- |
|  |  | **Most advanced finding**  **% (95% confidence interval)** | | | |
| **Sex** |  | **No neoplasm** | **Non-advanced adenoma** | **Advanced adenoma** | **Preclinical colorectal cancer** |
| Men | 50-54 | 69.4 (69.3 - 69.5) | 22.9 (22.8 – 23.0) | 7.1 (7 - 7.1) | 0.6 (0.6 - 0.6) |
|  | 55-59 | 69.4 (69.3 - 69.5) | 22.9 (22.8 - 23.1) | 7.1 (7 - 7.1) | 0.6 (0.6 - 0.7) |
|  | 60-64 | 65.6 (65.4 - 65.7) | 24.5 (24.4 - 24.7) | 8.8 (8.8 - 8.9) | 1.1 (1.0 - 1.1) |
|  | 65-69 | 62.6 (62.5 - 62.8) | 26.0 (25.8 - 26.2) | 9.9 (9.8 – 10.0) | 1.5 (1.4 - 1.5) |
|  | 70-74 | 60.2 (59.9 - 60.5) | 27.1 (26.8 - 27.3) | 10.6 (10.5 - 10.8) | 2.1 (2.0 - 2.2) |
| Women | 50-54 | 82.0 (81.9 - 82.1) | 13.9 (13.8 – 14.0) | 3.8 (3.7 - 3.9) | 0.3 (0.3 - 0.3) |
|  | 55-59 | 82.0 (81.9 - 82.2) | 13.9 (13.7 – 14.0) | 3.8 (3.7 - 3.9) | 0.3 (0.3 - 0.3) |
|  | 60-64 | 79.1 (78.9 - 79.2) | 15.5 (15.3 - 15.6) | 4.9 (4.9 – 5.0) | 0.5 (0.5 - 0.6) |
|  | 65-69 | 76.5 (76.3 - 76.7) | 17.1 (16.9 - 17.2) | 5.7 (5.6 - 5.8) | 0.7 (0.7 - 0.8) |
|  | 70-74 | 73.6 (73.4 - 73.9) | 18.7 (18.4 - 18.9) | 6.5 (6.4 - 6.7) | 1.2 (1.1 - 1.2) |
| ^1^ Estimates based on the German screening colonoscopy registry. Extracted and recalculated from reference [5] | | | | | |
| **A2. Calibrated to Italian population. Proportions of no neoplasm, non-advanced adenoma, advanced adenoma and preclinical CRC at the beginning of simulation**^1^ | | | | | |
|  |  | **Most advanced finding**  **% (95% confidence interval)** | | | |
| **Sex** |  | **No neoplasm** | **Non-advanced adenoma** | **Advanced adenoma** | **Preclinical colorectal cancer** |
| Men | 50-54 | 71.4 (71.3 - 71.5) | 21.4 (21.3 - 21.5) | 6.6 (6.5 - 6.6) | 0.6 (0.6 - 0.6) |
|  | 55-59 | 71.4 (71.3 - 71.5) | 21.4 (21.3 - 21.5) | 6.6 (6.5 - 6.6) | 0.6 (0.6 - 0.6) |
|  | 60-64 | 67.8 (67.7 – 68.0) | 22.9 (22.8 – 23.0) | 8.3 (8.2 - 8.3) | 1.0 (1.0 – 1.0) |
|  | 65-69 | 65.1 (64.9 - 65.3) | 24.3 (24.2 - 24.4) | 9.2 (9.2 - 9.3) | 1.4 (1.3 - 1.4) |
|  | 70-74 | 62.8 (62.6 – 63.0) | 25.3 (25.1 - 25.4) | 9.9 (9.8 - 10) | 2.0 (1.9 – 2.0) |
| Women | 50-54 | 83.1 (83.0 - 83.2) | 13.1 (13.0 - 13.1) | 3.6 (3.5 - 3.6) | 0.3 (0.3 - 0.3) |
|  | 55-59 | 83.1 (83.0 - 83.2) | 13.1 (13.0 - 13.1) | 3.6 (3.5 - 3.6) | 0.3 (0.3 - 0.3) |
|  | 60-64 | 80.3 (80.2 - 80.4) | 14.6 (14.5 - 14.7) | 4.7 (4.6 - 4.7) | 0.5 (0.5 - 0.5) |
|  | 65-69 | 77.9 (77.7 – 78.0) | 16.1 (16.0 - 16.2) | 5.4 (5.3 - 5.4) | 0.7 (0.7 - 0.7) |
|  | 70-74 | 75.2 (75.0 - 75.3) | 17.6 (17.5 - 17.7) | 6.1 (6.1 - 6.2) | 1.1 (1.1 - 1.1) |

^1^ Estimates based on the German screening colonoscopy registry. Extracted and recalculated from reference [5]. Calibrated to reflect lower incidence rates in the Italian vs the German population [25].

**Supplementary Table 1** Overview of model parameters (continued)

| **B. Sex- and age-specific annual transition rates between states^1^** | | | | | | | |  |
| --- | --- | --- | --- | --- | --- | --- | --- | --- |
|  |  |  | | **Annual transition rates**  **% (95% confidence interval)** | | | |  |
| **Sex** | **Age** | **No neoplasm to non-advanced adenoma^2^** | **No neoplasm to non-advanced adenoma^3^** | | **Non-advanced adenoma to advanced adenoma** | **Advanced adenoma to preclinical colorectal cancer** | **Preclinical colorectal cancer to clinical colorectal cancer** | **Preclinical colorectal cancer to colorectal cancer death** |
| Men | 50-54 | 3.1 (2.9 – 3.4) | 2.9 (2.7 – 3.2) | | 3.3 (2.8 – 3.9) | 2.6 (2.2 – 3.1) | 15.5 (14.9 – 16.6) | 1.5 (1.4 – 1.6) |
|  | 55-59 | 3.1 (2.9 – 3.4) | 2.9 (2.7 – 3.2) | | 3.3 (2.8 – 3.9) | 2.6 (2.2 – 3.1) | 15.5 (14.9 – 16.6) | 1.5 (1.4 – 1.6) |
|  | 60-64 | 3.1 (2.8 – 3.4) | 2.9 (2.6 – 3.2) | | 3.2 (2.6 – 3.7) | 3.1 (2.6 – 3.4) | 16.4 (15.7 – 17.4) | 1.6 (1.6 – 1.7) |
|  | 65-69 | 3.2 (2.9 – 3.4) | 3.0 (2.7 – 3.2) | | 3.2 (2.6 – 3.7) | 3.8 (3.4 – 4.3) | 18.2 (17.4 – 19.1) | 1.8 (1.7 – 1.9) |
|  | 70-74 | 2.9 (2.6 – 3.3) | 2.7 (2.4 – 3.1) | | 3.3 (2.6 – 4.0) | 5.1 (4.5 – 5.8) | 17.6 (16.8 – 18.5) | 1.7 (1.7 – 1.8) |
|  | 75-79 | 2.3 (1.8 – 2.9) | 2.1 (1.7 – 2.7) | | 3.0 (1.9 – 4.2) | 5.2 (4.2 – 6.2) | 17.3 (16.3 – 18.3) | 1.7 (1.6 – 1.8) |
|  | 80+ | 2.3 (1.8 – 2.9) | 2.1 (1.7 – 2.7) | | 3.0 (1.9 – 4.2) | 5.2 (4.2 – 6.2) | 15.7 (14.5 – 17.1) | 1.6 (1.4 –1.7) |
| Women | 50-54 | 1.8 (1.7 – 2.0) | 1.7 (1.6 – 1.9) | | 3.2 (2.6 – 3.8) | 2.5 (2.0 – 2.9) | 18.2 (16.8 – 19.7) | 1.9 (1.8 – 2.1) |
|  | 55-59 | 1.8 (1.7 – 2.0) | 1.7 (1.6 – 1.9) | | 3.2 (2.6 – 3.8) | 2.5 (2.0 – 2.9) | 18.2 (16.8 – 19.7) | 1.9 (1.8 – 2.1) |
|  | 60-64 | 2.0 (1.8 – 2.2) | 1.9 (1.7 – 2.1) | | 2.9 (2.2 – 3.4) | 2.7 (2.2 – 3.2) | 19.1 (17.8 – 20.3) | 2.0 (1.9 – 2.1) |
|  | 65-69 | 2.1 (1.9 – 2.3) | 2.0 (1.8 – 2.2) | | 2.9 (2.3 – 3.5) | 3.8 (3.3 – 4.3) | 18.7 (17.7 – 19.7) | 2.0 (1.9 – 2.1) |
|  | 70-74 | 2.0 (1.7 – 2.2) | 1.9 (1.6 – 2.1) | | 3.8 (3.0 – 4.6) | 5.0 (4.2 – 5.7) | 17.8 (16.8 – 18.9) | 1.9 (1.8 – 2.0) |
|  | 75-79 | 1.6 (1.1 – 2.0) | 1.5 (1.0 – 1.9) | | 3.0 (1.7 – 4.4) | 5.6 (4.4 – 6.8) | 16.5 (15.5 – 17.7) | 1.7 (1.6 – 1.9) |
|  | 80+ | 1.6 (1.1 – 2.0) | 1.5 (1.0 – 1.9) | | 3.0 (1.7 – 4.4) | 5.6 (4.4 – 6.8) | 14.9 (13.9 – 16.1) | 1.6 (1.4 – 1.7) |

^1^ Estimates extracted and recalculated from references [15–17]

^2^ Core model

^3^ Calibrated to reflect lower incidence rates in the Italian vs the German population [25].

**Supplementary Table 1** Overview of model parameters (continued)

| **C. Diagnostic performance parameters** | | | | | |
| --- | --- | --- | --- | --- | --- |
|  |  | **Performance (%)** | | | |
| **Test (sex)** | **Parameter** | **No neoplasm** | **Non-advanced adenoma** | **Advanced adenoma** | **Preclinical colorectal cancer** |
| Sigmoidoscopy (men) **³** | Sensitivity | - | 31.2 | 61.3 | 61.3 |
|  | Specificity | 100.0 | - | - | - |
| Sigmoidoscopy (women) **³** | Sensitivity | - | 30.8 | 58.5 | 58.5 |
|  | Specificity | 100.0 | - | - | - |
| Colonoscopy (both sexes) ^4^ | Sensitivity | - | 75.0 | 95.0 | 95.0 |
|  | Specificity | 100.0 | - | - | - |
| ^3^ Diagnostic performance for the entire colon. Estimated based on data from the BLITZ study [26].  ^4^ Estimates based on references [12,13] | | | | | |

#### **Supplementary Table 2.** Annual CRC-specific mortality rates of CRC patients by mode of cancer detection^1^

|  | **Annual CRC-specific mortality rates (%)** | | | |
| --- | --- | --- | --- | --- |
| **Year after diagnosis** | **Screening colonoscopy– detected cases** | | **Symptom-detected cases** | |
|  | **Men** | **Women** | **Men** | **Women** |
| 1 | 4.6 | 3.7 | 19.7 | 20.6 |
| 2 | 2.2 | 1.9 | 9.3 | 10.7 |
| 3 | 2.1 | 1.3 | 8.8 | 7.4 |
| 4 | 1.5 | 0.9 | 6.3 | 4.8 |
| 5 | 1.2 | 0.6 | 5.0 | 3.3 |
| 6 | 0.8 | 0.3 | 3.5 | 1.7 |
| 7 | 0.4 | 0.3 | 1.8 | 1.8 |
| 8 | 0.4 | 0.3 | 1.9 | 1.8 |
| 9 | 0.4 | 0.0 | 1.9 | 0.0 |
| 10 | 0.0 | 0.0 | 0.0 | 0.0 |

^1^ Estimates extracted from references [8,9]

CRC, Colorectal cancer.

#### **Supplementary Table 3.** Sex- and age-specific general mortality rates

|  | **General mortality rates from age to age +1 (%)** | | | |
| --- | --- | --- | --- | --- |
|  | **Core model (German population)** ^1^ | | **Calibrated to Italian population**^2^ | |
| **Age** | **Men** | **Women** | **Men** | **Women** |
| 50 | 0.4 | 0.2 | 0.3 | 0.2 |
| 51 | 0.4 | 0.2 | 0.3 | 0.2 |
| 52 | 0.5 | 0.3 | 0.3 | 0.2 |
| 53 | 0.6 | 0.3 | 0.4 | 0.2 |
| 54 | 0.6 | 0.3 | 0.4 | 0.2 |
| 55 | 0.7 | 0.4 | 0.5 | 0.3 |
| 56 | 0.7 | 0.4 | 0.5 | 0.3 |
| 57 | 0.8 | 0.4 | 0.6 | 0.3 |
| 58 | 0.9 | 0.4 | 0.6 | 0.3 |
| 59 | 1.0 | 0.5 | 0.7 | 0.4 |
| 60 | 1.0 | 0.5 | 0.8 | 0.4 |
| 61 | 1.1 | 0.6 | 0.8 | 0.4 |
| 62 | 1.2 | 0.6 | 0.9 | 0.5 |
| 63 | 1.3 | 0.7 | 1.0 | 0.5 |
| 64 | 1.4 | 0.7 | 1.1 | 0.5 |
| 65 | 1.5 | 0.8 | 1.2 | 0.6 |
| 66 | 1.7 | 0.9 | 1.3 | 0.7 |
| 67 | 1.8 | 0.9 | 1.5 | 0.8 |
| 68 | 1.9 | 1.0 | 1.6 | 0.8 |
| 69 | 2.1 | 1.1 | 1.8 | 0.9 |
| 70 | 2.2 | 1.2 | 2.0 | 1.0 |
| 71 | 2.4 | 1.3 | 2.1 | 1.1 |
| 72 | 2.7 | 1.4 | 2.3 | 1.2 |
| 73 | 3.0 | 1.6 | 2.6 | 1.4 |
| 74 | 3.3 | 1.8 | 3.0 | 1.6 |
| 75 | 3.7 | 2.1 | 3.3 | 1.8 |
| 76 | 4.1 | 2.4 | 3.7 | 2,1 |
| 77 | 4.6 | 2.7 | 4.1 | 2,4 |
| 78 | 5.2 | 3.1 | 4.7 | 2,7 |
| 79 | 5.8 | 3.6 | 5.2 | 3.1 |

*Continued on next page*

**Supplementary Table 3.** Sex- and age-specific general mortality rates *(continued)*

|  | **General mortality rates from age to age +1 (%)** | | | |
| --- | --- | --- | --- | --- |
|  | **Core model (German population)** ^1^ | | **Calibrated to Italian population**^2^ | |
| **Age** | **Men** | **Women** | **Men** | **Women** |
| 80 | 6.5 | 4.1 | 6.0 | 3.5 |
| 81 | 7.2 | 4.7 | 6.8 | 4.1 |
| 82 | 8.0 | 5.4 | 7.6 | 4.7 |
| 83 | 8.9 | 6.2 | 8.5 | 5.5 |
| 84 | 9.9 | 7.1 | 9.6 | 6.3 |
| 85 | 11.1 | 8.2 | 10.6 | 7.2 |
| 86 | 12.3 | 9.3 | 12.0 | 8.3 |
| 87 | 13.7 | 10.7 | 13.3 | 9.5 |
| 88 | 15.3 | 12.1 | 14.5 | 10.6 |
| 89 | 16.9 | 13.7 | 15.4 | 11.6 |
| 90 | 18.7 | 15.4 | 16.9 | 12.9 |
| 91 | 20.7 | 17.2 | 18.8 | 14.7 |
| 92 | 22.7 | 19.1 | 22.1 | 17.7 |
| 93 | 24.8 | 21.1 | 25.3 | 20.5 |
| 94 | 27.0 | 23.2 | 27.6 | 22.7 |
| 95 | 29.1 | 25.3 | 29.6 | 24.4 |
| 96 | 31.2 | 27.4 | 30.4 | 25.6 |
| 97 | 33.2 | 29.6 | 31.9 | 27.2 |
| 98 | 35.1 | 31.7 | 33.7 | 29.2 |
| 99 | 37.2 | 34.0 | 35.9 | 31.7 |
| 100 | 39.2 | 36.2 | 39.1 | 34.9 |

^1^Estimates were extracted from German population life tables 2010/2012 (reference [20])

^2^Estimates were extracted from Italian population life tables 2010/2012 (reference [34])

#### **Supplementary Table 4. Standardized colorectal cancer incidence rate in Germany and Italy, 2000-2004**

|  |  |  |  | **Standardized incidence rate ^1^** | |  | **Ratio**  **(Italy / Germany)** | **Discount (Italy v Germany)** |
| --- | --- | --- | --- | --- | --- | --- | --- | --- |
| **Sex** |  | **Year** |  | **Germany** | **Italy** |  |  |  |
| men |  | 2000 |  | 65.29 | 60.26 |  | 0.92 | 0.08 |
|  |  | 2001 |  | 64.94 | 65.38 |  | 1.01 | -0.01 |
|  |  | 2002 |  | 70.46 | 66.15 |  | 0.94 | 0.06 |
|  |  | 2003 |  | 68.35 | 59.15 |  | 0.87 | 0.13 |
|  |  | 2004 |  | 68.33 | 64.06 |  | 0.94 | 0.06 |
|  |  |  |  |  |  |  | *5-year average:* | *0.06* |
| women |  | 2000 |  | 42.67 | 45.10 |  | 1.06 | -0.06 |
|  |  | 2001 |  | 43.30 | 40.37 |  | 0.93 | 0.07 |
|  |  | 2002 |  | 44.67 | 37.14 |  | 0.83 | 0.17 |
|  |  | 2003 |  | 45.01 | 40.99 |  | 0.91 | 0.09 |
|  |  | 2004 |  | 44.10 | 43.22 |  | 0.98 | 0.02 |
|  |  |  |  |  |  |  | *5-year average:* | *0.06* |

1 Extracted from reference [25]

#### **Supplementary Table 5. Apparent and true incidence rate ratios, risk reduction and underestimation of risk reduction in the simulated SCORE over time, stratified by sex**

|  | **Year of follow-up** | **IRR** | |  | **Risk reduction** | |  | **Underestimation  of risk reduction** | |
| --- | --- | --- | --- | --- | --- | --- | --- | --- | --- |
| **Sex** |  | **Apparent** | **True^1^** |  | **Apparent** | **True** |  | **Absolute** | **Relative** |
| **Intention-to-screen analysis** | | | | | | | | | |
| **Men** | 5 | 0.97 | 0.66 |  | 3% | 34% |  | 31 % units | 91% |
|  | 8 | 0.84 | 0.68 |  | 16% | 32% |  | 16 % units | 50% |
|  | 11 | 0.81 | 0.71 |  | 19% | 29% |  | 10 % units | 34% |
|  | 15 | 0.79 | 0.74 |  | 21% | 26% |  | 5 % units | 19% |
| **Women** | 5 | 0.94 | 0.68 |  | 6% | 32% |  | 26 % units | 81% |
|  | 8 | 0.84 | 0.70 |  | 16% | 30% |  | 14 % units | 47% |
|  | 11 | 0.81 | 0.73 |  | 19% | 27% |  | 8 % units | 30% |
|  | 15 | 0.80 | 0.76 |  | 20% | 24% |  | 4 % units | 17% |
| **Per-protocol analysis** | | | | | | | | | |
| **Men** | 5 | 0.95 | 0.41 |  | 5% | 59% |  | 54 % units | 92% |
|  | 8 | 0.73 | 0.45 |  | 27% | 55% |  | 28 % units | 51% |
|  | 11 | 0.66 | 0.50 |  | 34% | 50% |  | 16 % units | 32% |
|  | 15 | 0.64 | 0.55 |  | 36% | 45% |  | 9 % units | 20% |
| **Women** | 5 | 0.90 | 0.44 |  | 10% | 56% |  | 46 % units | 82% |
|  | 8 | 0.72 | 0.48 |  | 28% | 52% |  | 24 % units | 46% |
|  | 11 | 0.67 | 0.53 |  | 33% | 47% |  | 14 % units | 30% |
|  | 15 | 0.66 | 0.59 |  | 34% | 41% |  | 7 % units | 17% |

^1^ ‘true’ incidence rate ratio: excluding prevalent cancers at baseline not preventable by screening.

IRR, incidence rate ratio

#### **Supplementary Table 6. Apparent and true incidence rate ratios, risk reduction and underestimation of risk reduction in the simulated SCORE over time, stratified by age**

|  | **Year of follow-up** | **IRR** | |  | **Risk reduction** | |  | **Underestimation  of risk reduction** | |
| --- | --- | --- | --- | --- | --- | --- | --- | --- | --- |
| **Age** |  | **Apparent** | **True**^1^ |  | **Absolute** | **Relative** |  | **Absolute** | **Relative** |
| **Intention-to-screen analysis** | | | | | | | | | |
| **55-59** | 5 | 0.96 | 0.67 |  | 4% | 33% |  | 29 % units | 88% |
|  | 8 | 0.83 | 0.69 |  | 17% | 31% |  | 14 % units | 45% |
|  | 11 | 0.80 | 0.72 |  | 20% | 28% |  | 8 % units | 29% |
|  | 15 | 0.79 | 0.75 |  | 21% | 25% |  | 4 % units | 16% |
| **60-64** | 5 | 0.95 | 0.66 |  | 5% | 34% |  | 29 % units | 85% |
|  | 8 | 0.84 | 0.68 |  | 16% | 32% |  | 16 % units | 50% |
|  | 11 | 0.80 | 0.71 |  | 20% | 29% |  | 9 % units | 31% |
|  | 15 | 0.80 | 0.74 |  | 20% | 26% |  | 6 % units | 23% |
| **Per-protocol analysis** | | | | | | | | | |
| **55-59** | 5 | 0.93 | 0.42 |  | 7% | 58% |  | 51 % units | 88% |
|  | 8 | 0.71 | 0.47 |  | 29% | 53% |  | 24 % units | 45% |
|  | 11 | 0.65 | 0.51 |  | 35% | 49% |  | 14 % units | 29% |
|  | 15 | 0.64 | 0.57 |  | 36% | 43% |  | 7 % units | 16% |
| **60-64** | 5 | 0.92 | 0.42 |  | 8% | 58% |  | 50 % units | 86% |
|  | 8 | 0.72 | 0.45 |  | 28% | 55% |  | 27 % units | 49% |
|  | 11 | 0.66 | 0.50 |  | 34% | 50% |  | 16 % units | 32% |
|  | 15 | 0.65 | 0.56 |  | 35% | 44% |  | 9 % units | 20% |

^1^ ‘true’ incidence rate ratio: excluding prevalent preclinical cancers at baseline not preventable by screening.

IRR, incidence rate ratio

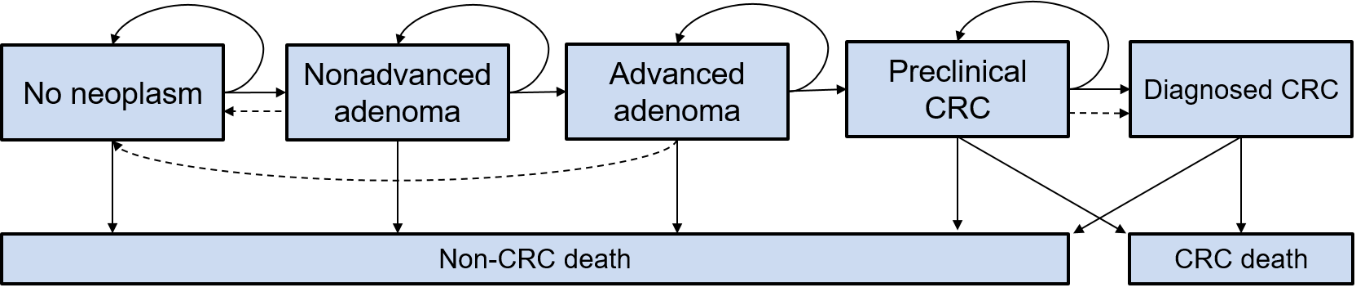

#### **Supplementary Figure 1. Schematic illustration of the Colorectal Cancer Multistate Simulation Model (COSIMO)**

Solid lines represent the progression of colorectal disease through the adenoma-carcinoma sequence in the absence of screening; dashed lines show the movement between states because of the detection and removal of adenomas and the detection of asymptomatic CRC at screening.

CRC, Colorectal cancer.

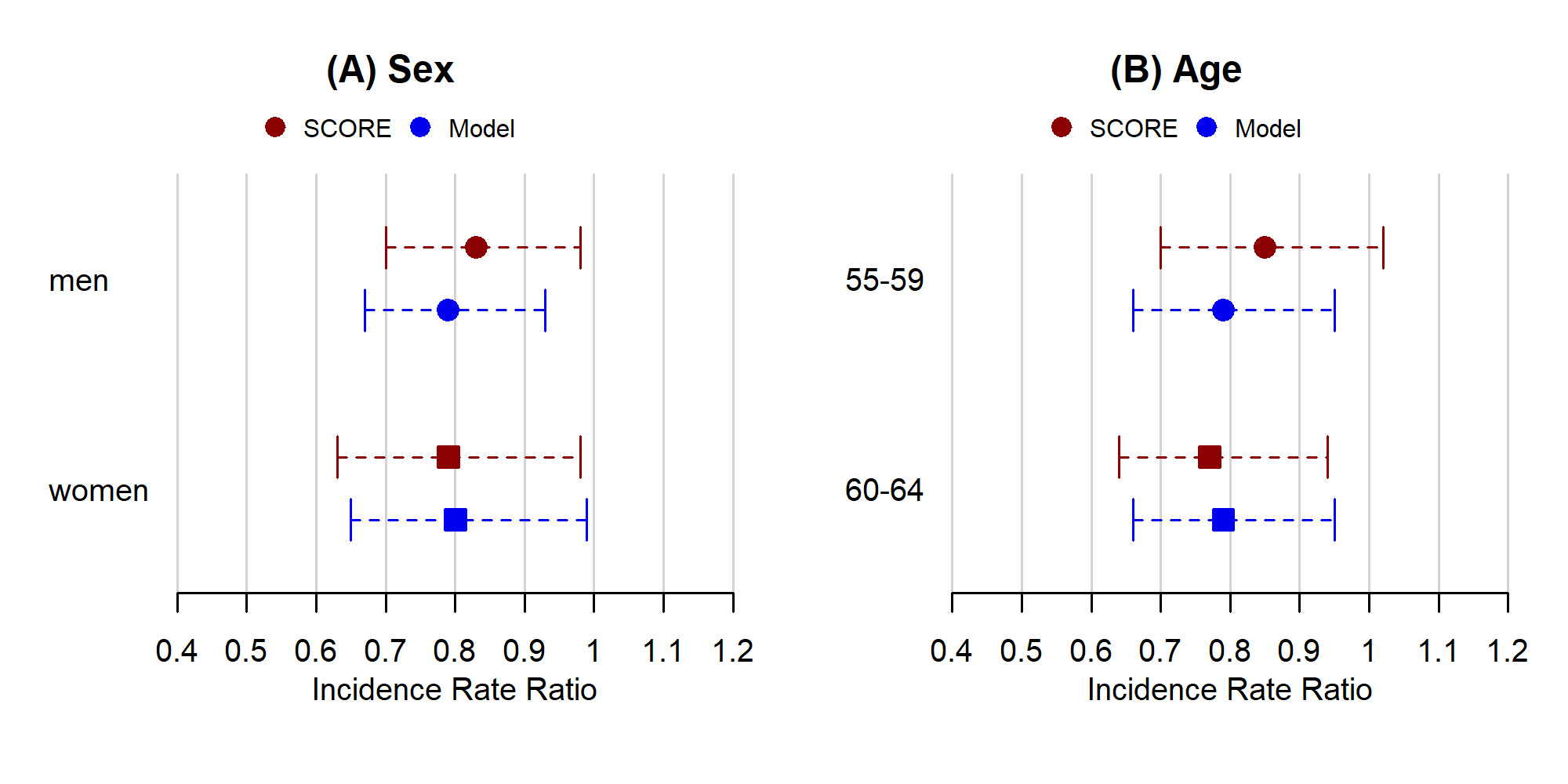

#### **Supplementary Figure 2. Colorectal cancer incidence rate ratios (‘apparent’) in the actual as well as the simulated SCORE trial by sex and age groups after 15 years of follow-up**

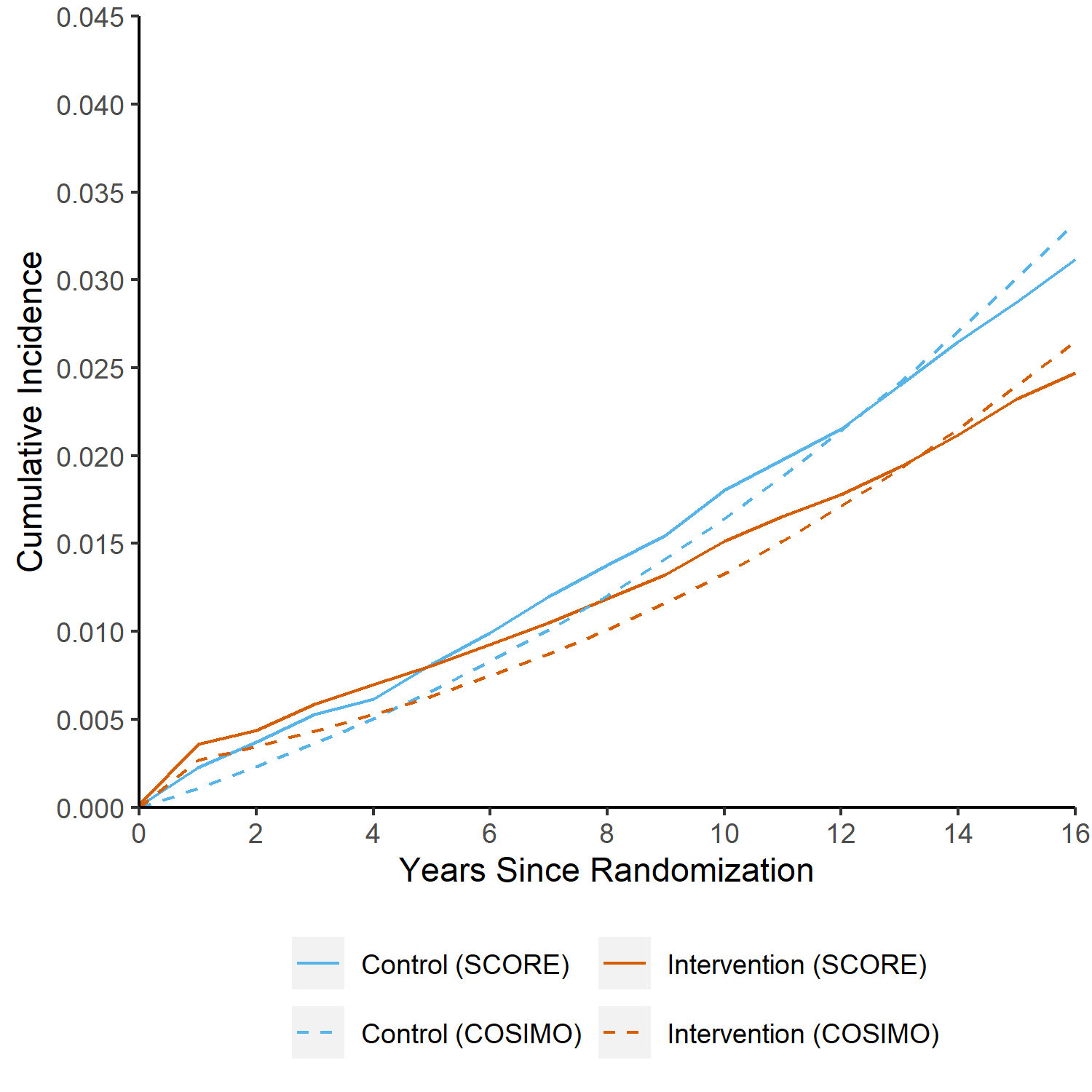

#### **Supplementary Figure 3. Cumulative incidence of colorectal cancer by time from randomization in the actual* and simulated SCORE trial (overall trial population, intention-to-screen analysis)**

* approximated from reference [24]

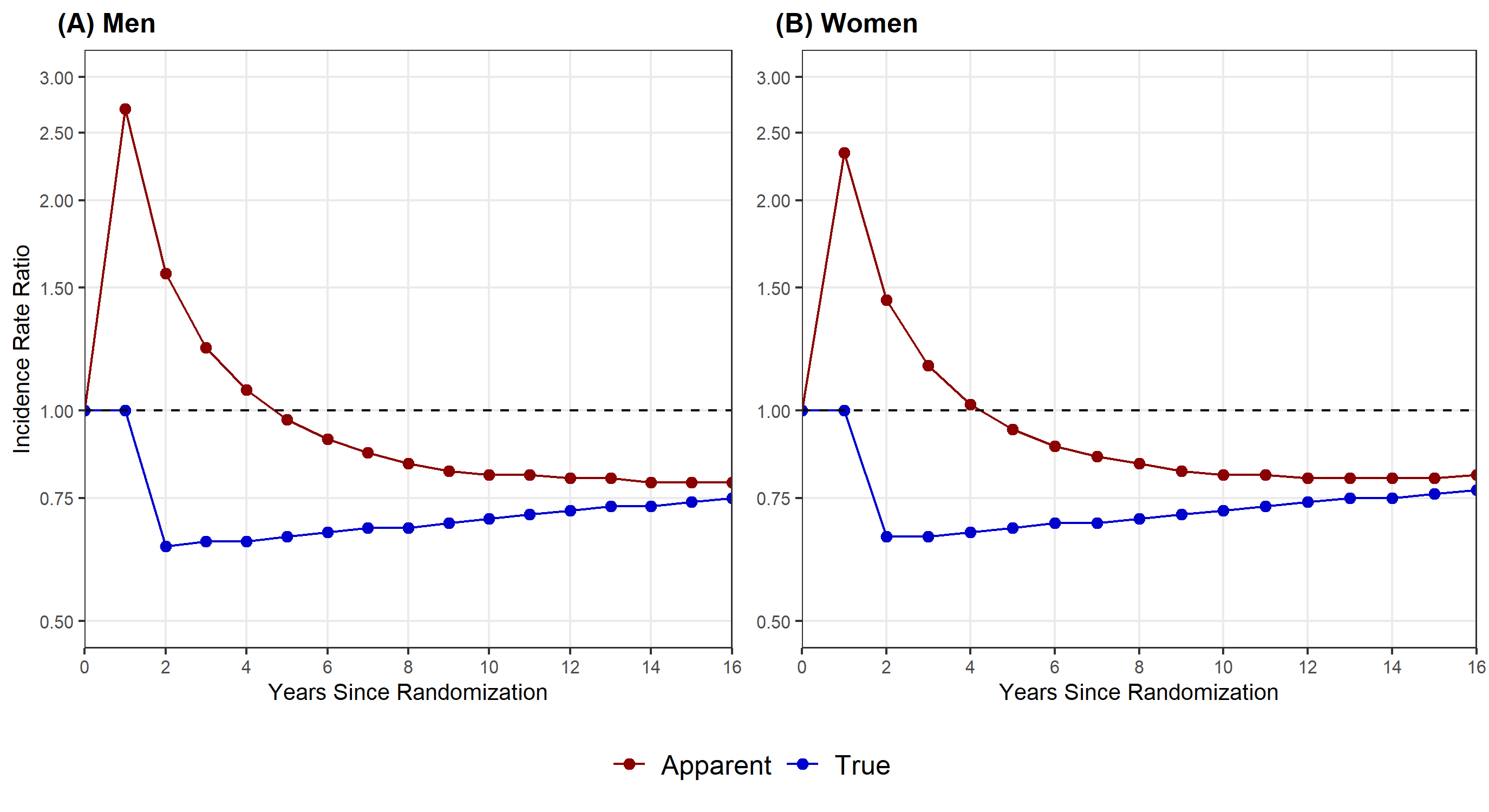

#### **Supplementary Figure 4. Apparent and true incidence rate ratios (intention-to-screen analysis) in the simulated SCORE trial over time, stratified by sex**

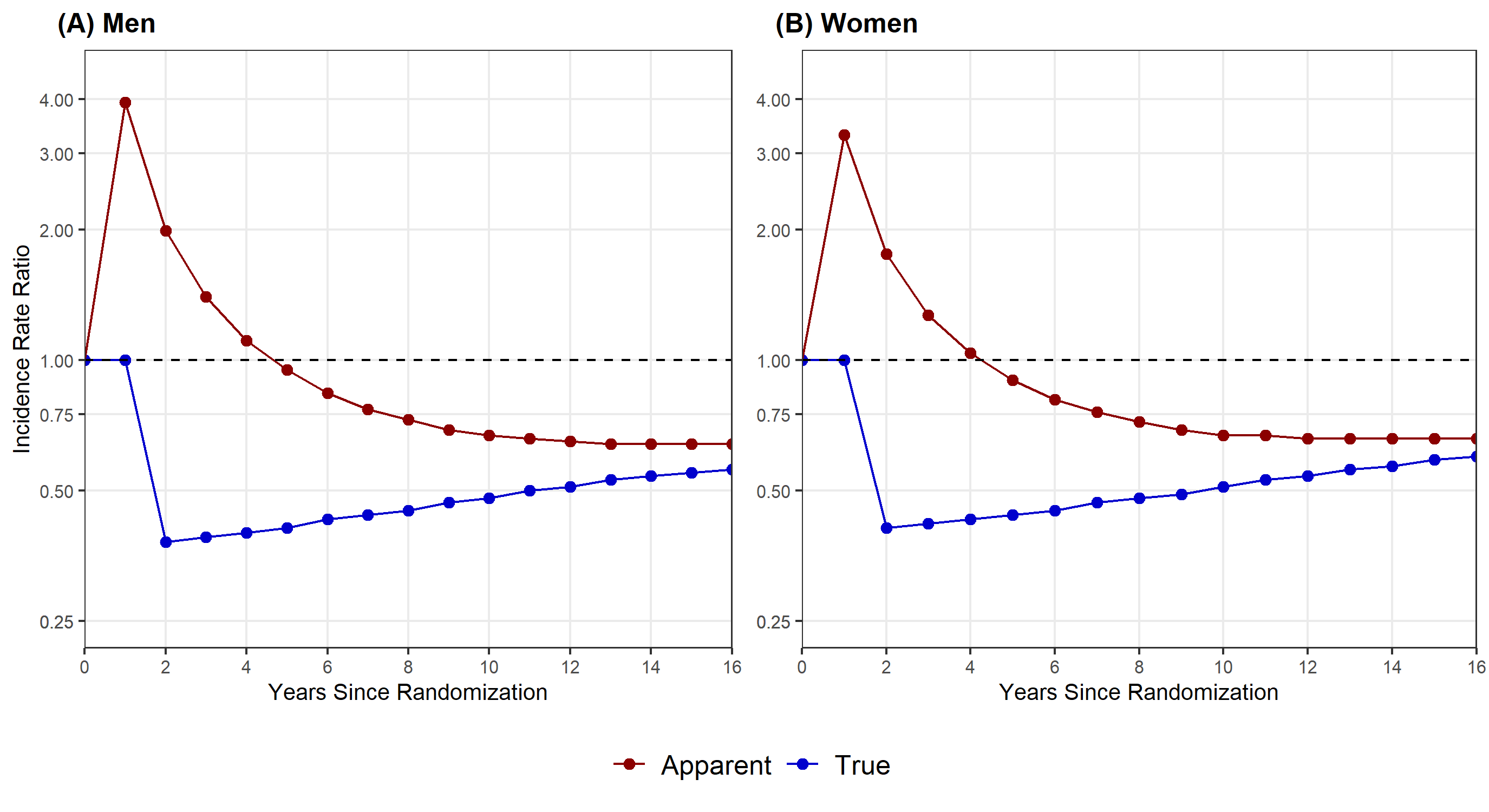

#### **Supplementary Figure 5. Apparent and true incidence rate ratios (per-protocol analysis) in the simulated SCORE trial over time, stratified by sex**

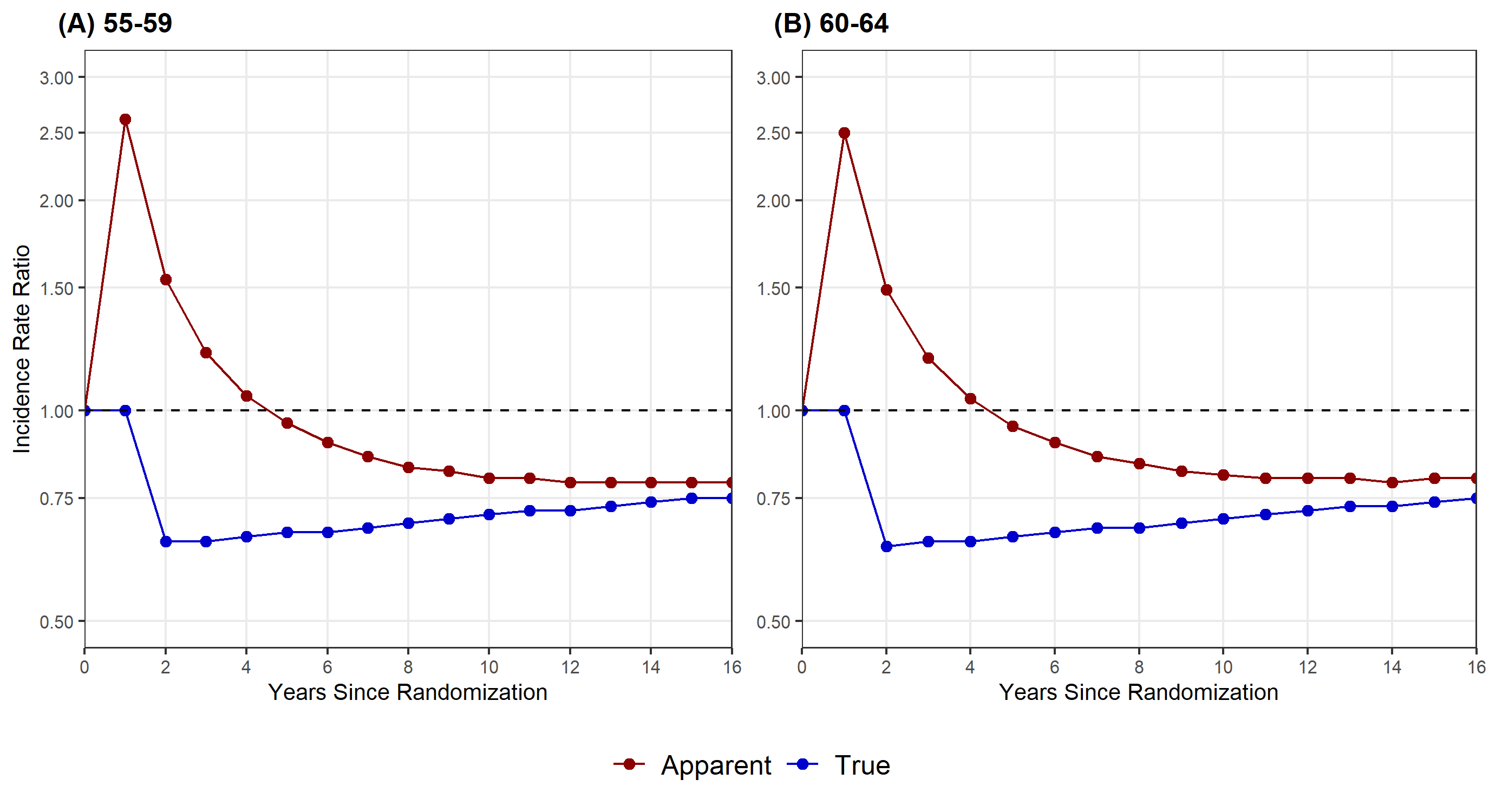

#### **Supplementary Figure 6. Apparent and true incidence rate ratios (intention-to-screen analysis) in the simulated SCORE trial over time, stratified by age**

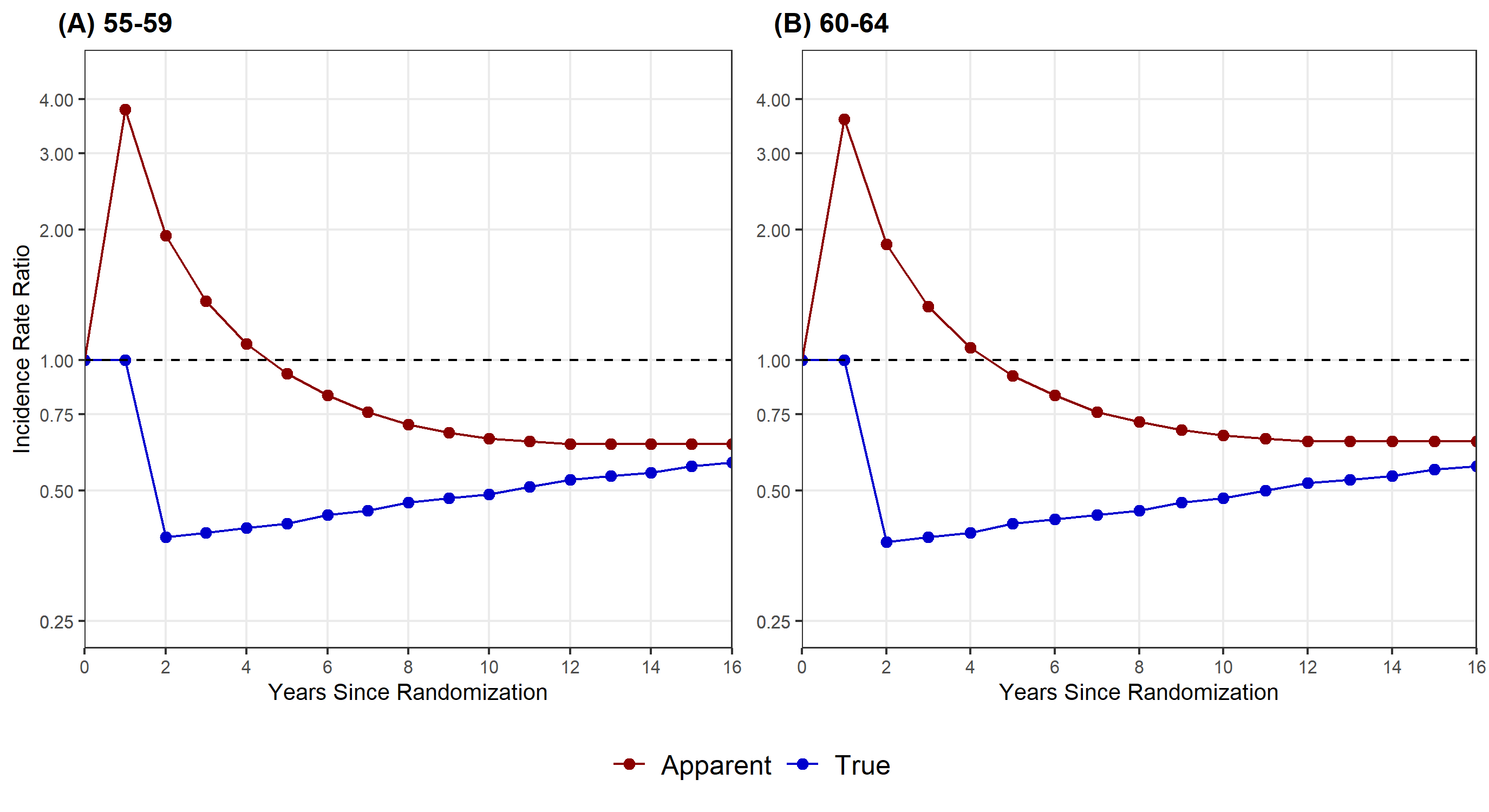

#### **Supplementary Figure 7. Apparent and true incidence rate ratios (per-protocol analysis) in the simulated SCORE trial over time, stratified by age**
